# Subjective and Objective Measures of Cognitive Function are Correlated in Persons with Post-COVID-19 Condition: A Secondary Analysis of a Randomized Controlled Trial

**DOI:** 10.1101/2024.03.20.24304410

**Authors:** Angela T.H. Kwan, Moiz Lakhani, Gia Han Le, Gurkaran Singh, Kayla M. Teopiz, Felicia Ceban, Charnjit S. Nijjar, Shakila Meshkat, Sebastian Badulescu, Roger Ho, Taeho Greg Rhee, Joshua D. Di Vincenzo, Hartej Gill, Roger S. McIntyre

## Abstract

**Background:** It remains unclear whether subjective and objective measures of cognitive function in Post COVID-19 Condition (PCC) are correlated. The extent of correlation has mechanistic and clinical implications.

**Methods:** This post-hoc analysis of a randomized, double-blind, placebo-controlled clinical trial contains baseline data of subjective and objective measures of cognition in a rigorously characterized cohort living with PCC. Herein, we evaluated the association between subjective and objective condition function, as measured by the Perceived Deficits Questionnaire, 20-item (PDQ-20) and the Digit Symbol Substitution Test (DSST) and Trails Making Test (TMT)-A/B, respectively.

**Results:** A total of 152 participants comprised the baseline sample. Due to missing data, our statistical analyses included 150 for self-reported PDQ-20, 147 individuals for combined DSST-measured cognitive function (composite z-score of the Pen/Paper plus Online CogState Version, *N*_*combinedDSST*_), 71 for in-person DSST-measured cognitive function (Pen/Paper Version), 70 for TMT-A-measured cognitive function, and 70 for TMT-B-measured cognitive function. After adjusting for age, sex, and education, PDQ-20 was significantly correlated with pen-and-paper DSST (β = -0.003, *p* = 0.002) and TMT-B (β = 0.003, *p* = 0.008) scores, but not with TMT-A scores (β = -0.001, *p* = 0.751).

**Conclusions:** Overall, a statistically significant correlation was observed between subjective and objective cognitive functions. Clinicians providing care for individuals with PCC who have subjective cognitive function complaints may consider taking a measurement-based approach to cognition at the point of care that focuses exclusively on patient-reported measures.

## INTRODUCTION

Post COVID-19 Condition (PCC) is a common, persistent, and debilitating phenomenon defined by the World Health Organization (WHO) as occurring three months after a confirmed coronavirus disease 2019 (COVID-19) infection despite the resolution of the acute infection and is unexplainable by an alternative diagnosis.^1,2^ Approximately 10-20% of individuals infected with COVID-19 meet the criteria for PCC.^3,4^ Around 22% of people who suffered from PCC reported cognitive impairment.^1^ Symptoms of cognitive impairment collectively described as *“brain fog”* (e.g., memory impairment) manifest independently of mental health conditions and have been ascribed to PCC.^1,5–8^ Extant evidence and WHO consensus describe neurological manifestations of PCC as one of the most common persistent symptoms. Occurring in over a fifth of individuals, clinical presentations can involve attention, executive functioning, language, memory, and processing speed.^1,3,9^ Cognitive impairments have been reported to have a significant negative impact on working ability and health-related quality of life.^10,11^

The relevance of cognitive deficits in PCC is underscored by data indicating that they are a principal quality of life detractor and mediator of functional impairment.^12,13^ Extant evidence indicates that both subjective and objective cognitive deficits (subjective self-reports on cognitive abilities versus objective performance on neuropsychological tests) are apparent in persons with PCC.^14,15^ However, the presence and magnitude of cognitive deficits reported are mixed, likely reflecting differences in methodological approaches and the heterogeneity of PCC. Studies conducted in other medical conditions [e.g., Major Depressive Disorder (MDD), Diabetes Mellitus] indicated that subjective and objective cognitive functions are often not correlated.^14,16–20^ For patients with MDD, the discrepancy between objective and subjective cognitive measures has been hypothesized to be as result of greater depressive symptoms, intelligence quotient and executive function that leads to underestimation of attentional and memory abilities.^20^

Individuals living with PCC often express subjective cognitive issues as their main concern motivating the clinical visit.^21^ What remains insufficiently characterized is the extent to which subjective cognitive complaints in persons living with PCC correlate with objective cognitive function. Further data evaluating this relationship regarding PCC has clinical, conceptual, and therapeutic implications. For example, it is important to determine if the neurobiological substrates subserving subjective cognitive performance in PCC overlap with the neurological substrates subserving objective cognitive function. Separately, for persons presenting clinically with PCC and subjective cognitive complaints, the extent that subjective cognitive complaints can be taken as prima facie evidence of objective cognitive impairment would be informed by the degree of correlation. This would also inform whether supplementary objective cognitive testing is required. Thus, if a high correlation between subjective and objective cognitive functions exists, then that would imply that a measurement-based approach to cognition at point-of-care could depend exclusively on patient reported measures. Herein, we sought to determine the association between subjective and objective cognitive functions in a well-characterized cohort of adults between 18-65 meeting the WHO criteria for PCC.

## MATERIALS AND METHODS

### Study Design and Participants

This is a post-hoc analysis of a randomized, double-blind, placebo-controlled clinical trial evaluating vortioxetine for the treatment of cognitive deficits in adults with PCC. The data and methodology are obtained from a primary study which is published elsewhere.^22^ The protocol of the primary trial was registered on Clinicaltrials.gov (identifier number: NCT05047952) and approved by Advarra, which is a local research ethics board that complies with Health Canada regulations (IRB #00000971).

The primary trial recruited participants from Canada and was conducted in accordance with the principles of Good Clinical Practice (ICH, 1996) and the Declaration of Helsinki (WMA, 2008).^23,24^ Participant recruitment occurred from November 2021 to January 2023 via media advertisements (e.g., Facebook, Twitter, Instagram, print) or referrals by medical practitioners.

Eligible participants were ≥ 18 years of age, resided in Canada, were able to provide documentation of a positive severe acute respiratory syndrome coronavirus 2 (SARS□CoV□2) test (i.e., antigen, serology, and/or PCR) as well as a documented history of experiencing acute-COVID-19 symptoms post-SARS-CoV-2 infection occurring within 3 months after the acute COVID-19 infection and persisting for at least 2 months post-acute infection (i.e., meeting WHO criteria for PCC). In the absence of a prior positive SARS-CoV-2 test, participants with a confirmation of a probable SARS-CoV-2 infection (i.e., a presumptive prior acute-COVID-19 case) from a healthcare provider or a clinical diagnosis from the study physician were deemed eligible. The pre-screening of interested individuals was conducted by trained personnel and followed the trial’s inclusion and exclusion criteria **(Supplementary Materials, Table S1)**.

Additionally, eligible subjects were required to report subjective cognitive impairment, measured by the 20-item Perceived Deficits Questionnaire (PDQ-20). The study physician advised any participant that was currently taking other antidepressants to discontinue the antidepressant use for at least 2-4 weeks prior to their baseline visit. The informed consent forms conveyed to the participants that the simultaneous use of two antidepressants would be considered investigational and that the safety/efficacy profiles are unknown.

Participants who met study inclusion criteria and were able to provide written informed consent at the time of screening and baseline were eligible for inclusion in the study.

### Procedures

A total of 149 eligible participants were randomized (1:1) in a group that received either vortioxetine (5-20 mg/d) or placebo for 8 weeks of double-blind treatment. Within the vortioxetine group, the dose that participants received was dependent on their age; participants aged 18-65 years received 10 mg/d of vortioxetine in weeks 1 and 2, then from weeks 3 to 8 they received 20 mg/d. Participants aged 65+ years in the vortioxetine group received 5 mg/d of vortioxetine in weeks 1 and 2, then from weeks 3 to 8 they received 10 mg/d. Participants underwent assessment at the study site at baseline, and at weeks 2, 4 and 8.

Zoom and/or telephone meetings were also implemented to follow public health social distancing recommendations; this allowed visits to occur remotely and increased participant retention. The majority of visits with the study physician were conducted on a secure online platform (i.e., Ontario Telemedicine Network). Additionally, the participants were offered a follow-up safety visit between weeks 8 to 10. If a participant stopped the treatment course prematurely, they were scheduled for evaluation on the earliest possible date.

The medication capsules used in this trial were all indistinguishable in appearance. Additionally, participants randomized to treatment were instructed to take their assigned dose (i.e., 5-20 mg/day) orally at the same time each day.

### Outcome Measures

The primary outcome measure in the primary clinical trial was the effect of vortioxetine versus placebo on cognitive function as measured by the Digital Symbol Substitution Test (DSST) Pen/Paper Version and Online CogState Version as part of the CogState Online Cognitive Battery.^22^ Remote participants did not complete the pen/paper version of the DSST. The DSST was administered at baseline, and weeks 2 and 8.

Secondary outcome measures in the primary clinical trial included baseline-to-endpoint changes in the CogState Online Cognitive Battery, Trails Making Test (TMT)-A/B, and the 20-item Perceived Deficits Questionnaire (PDQ-20).^22^ The CogState Online Cognitive Battery and TMT-A/B were measured at baseline, and weeks 2 and 8. The PDQ-20 was measured as baseline and weeks 2, 4 and 8. A comprehensive list of all secondary outcomes in the primary trial are reported elsewhere.^22^

In the post-hoc analysis herein, we evaluated the association between subjective measures of cognition, as measured by the PDQ-20, and objective measures of cognition, as measured by the DSST and TMT-A/B in adults with PCC at baseline.

### Statistical Analysis

All statistical analyses were conducted using the IBM SPSS Statistics software, version 28.0.1.1 (15) with two-sided statistical significance set at α = 0.05. Descriptive statistics were written as frequency (%) for categorical variables; the mean [standard deviation (SD)] and median were reported for normally distributed numerical variables.

Of the 149 enrolled participants, 11 (7.4%) completed the Pen/Paper Version of the DSST only; 78 (52.3%) completed only the CogState Version, and 60 (40.3%) completed both the Pen/Paper and Online CogState Version. For participants that completed both DSST versions, performances on the Pen/Paper and Online CogState Version were highly and significantly correlated (r = 0.588, p < 0.001). Since not all participants completed both the Pen/Paper and Online CogState Versions, further analyses were performed using the combined DSST scores. The combined DSST scores were based on participants’ Online CogState DSST scores if they completed both Online CogState and Pen/Paper DSST. Participants’ Pen/Paper DSST scores were included in the combined DSST scores if the Online CogState DSST was not completed. For the assessment of subjective and objective DSST total scores at baseline, an intent-to-treat (ITT) analysis (i.e., including all randomized participants) was employed.

A generalized linear model (GLM) with a Poisson probability distribution was performed to examine the association between subjective (PDQ-20) and objective cognition (DSST and TMT-A/B). In cases where the data collected was not in the form of whole integers, a linear regression analysis was conducted. Additionally, the participants’ age, sex, and education were considered as covariates. The significance level was set at p < 0.05 to determine the statistical significance of the findings.

## RESULTS

### Patient Characteristics

Baseline sociodemographic and overall objective and subjective cognitive test results are described in **Table 1**. A total of 200 patients provided written informed consent; however, only data from 147 participants were used in baseline analysis due to missing screening/baseline data.

**Table 1.** Baseline characteristics of the intent-to-treat population (N = 147).

|  |  |  |  |  |  |  |  | Shapiro-Wilk |
| --- | --- | --- | --- | --- | --- | --- | --- | --- |
| Descriptives | N (%) | Mean (SD) | Std. Error Mean | Median | Minimum | Maximum | W | p |
| Age | 147 | 44.4 (12.2) | 1.01 | 44.0 | 20 | 73 | 0.983 | 0.060 |
| Sex | 147 |  |  |  |  |  |  |  |
| Female | 111 (75.5) |  |  |  |  |  |  |  |
| Male | 36 (24.5) |  |  |  |  |  |  |  |
| Education | 147 |  |  |  |  |  |  |  |
| < High School | 1 (0.68) |  |  |  |  |  |  |  |
| High School Graduate | 12 (8.2) |  |  |  |  |  |  |  |
| College/University Degree | 17 (11.6) |  |  |  |  |  |  |  |
| Associates Degree | 28 (19.1) |  |  |  |  |  |  |  |
| Bachelor's Degree | 61 (41.5) |  |  |  |  |  |  |  |
| Graduate Degree | 24 (16.3) |  |  |  |  |  |  |  |
| Professional Degree | 4 (2.7) |  |  |  |  |  |  |  |
| PDQ-20 | 146 | 68.7 (16.1) | 1.33 | 69.0 | 26 | 99 | 0.979 | 0.026 |
| Combined DSST | 147 | 48.0 (11.5) |  |  |  |  |  |  |
| Combined DSST Z-Score<sup>b</sup> | 147 | -0.233 (0.998) | 0.0823 | -0.199 | -3.17 | 1.89 | 0.988 | 0.245 |
| **Pen-and-Paper DSST** | 72 | 68.0 (15.2) | 1.80 | 70.0 | 22 | 101 | 0.990 | 0.825 |
| **TMT-A** | 71 | 29.6 (24.7) | 2.93 | 24.2 | 13.7 | 214.0 | 0.429 | < 0.001 |
| **TMT-B** | 71 | 55.3 (46.6) | 5.53 | 45.9 | 18.0 | 411.0 | 0.416 | < 0.001 |

a. Empty cells represent inapplicable information
b. Combined DSST z-score defined as Pen/Paper plus Online CogState Version.
Abbreviations: PDQ-20 = Perceived Deficits Questionnaire, 20-item; DSST = Digit Symbol Substitution Test, TMT-A = Trails Making Test-A, TMT-B = Trails Making Test-B
**Abbreviations:** DSST = Digit Symbol Substitution Test; TMT-A = Trail Making Test A; TMT-B = Trail Making Test B; and Perceived Deficits Questionnaire, 20-item (PDQ-20).

**Table 2.** Generalized linear model for the relationship between PDQ-20 and Pen/Paper DSST scores, after adjusting for age, sex, and education.

| Model | $\beta$ | Coefficients<br>Standard Error | 95% Confidence Interval | | *P*-value |
| --- | --- | --- | --- | --- | --- |
|  |  |  | Lower | Upper |  |
| **PDQ-20 Total** | -0.003 | 0.0009 | -0.005 | -0.001 | **0.002** |
| **Age** | -0.003 | 0.0011 | -0.005 | -0.001 | 0.003 |
| **Sex** | -0.015 | 0.0163 | -0.046 | 0.017 | 0.373 |
| **Education** | 0.009 | 0.0120 | -0.014 | 0.033 | 0.446 |

### Relationship Between PDQ-20 and DSST Performance in Persons with Post-COVID-19 Condition

A linear regression analysis was conducted on 149 patients’ baseline objective (DSST) and subjective (PDQ-20) cognitive test scores. Results from the linear regression analysis indicates that the overall regression, considering PDQ-20 and all covariates (i.e., age, sex, and education), is significant (r^2^ = 0.110, adjusted r^2^ = 0.085, df = 4, F = 4.385, *p* = 0.002) **(Table S2)**. In contrast, linear regression analysis indicates that the overall regression between all covariates and in-person DSST scores are not significant (r^2^ = 0.096, adjusted r^2^ = 0.042, df = 4, F = 1.758, *p* = 0.148) **(Table S3)**.

GLM analysis was subsequently conducted due to poor R squared results. The GLM analysis results indicate that PDQ-20 (β = -0.003, *p* = 0.002) and age (β = -0.003, *p* = 0.003) are significantly negatively correlated with in-person DSST scores. Consistently, a significant negative correlation exists between PDQ-20 scores and performance on the combined DSST, including Cogstate and pen-and-paper DSST (β = -0.002, *p* = 0.002) **(Table 3)**.

**Table 3.** Generalized linear model for the relationship between PDQ-20 and combined DSST (Pen/Paper and Online CogState), after adjusting for age, sex, and education.

| Model | $\beta$ | Coefficients<br>Standard Error | 95% Confidence Interval | | *P*-value |
| --- | --- | --- | --- | --- | --- |
|  |  |  | Lower | Upper |  |
| **Intercept** | 4.257 | 0.0932 | 4.074 | 4.439 | 0.000 |
| **PDQ-20 Total** | -0.002 | 0.0008 | -0.004 | -0.001 | **0.002** |
| **Age** | -0.006 | 0.0010 | -0.008 | -0.004 | < 0.001 |
| **Sex** | 0.005 | 0.0279 | -0.049 | 0.060 | 0.848 |
| **Education** | 0.007 | 0.0098 | -0.012 | 0.027 | 0.452 |

### Relationship Between PDQ-20 and TMT-A/B Performance in Persons with Post-COVID-19 Condition

In contrast to the aforementioned findings, the PDQ-20 is not significantly correlated with performance on the TMT-A (r^2^ = 0.005, adjusted r^2^ = -0.056, df = 4, F = 0.078, *p* = 0.989) and TMT-B (r^2^ = 0.003, adjusted r^2^ = -0.058, df = 4, F = 0.047, *p* = 0.996). Consistently, after adjusting for all covariates, PDQ-20 scores are not significantly correlated with performance on the TMT-A **(Table S4)** and TMT-B **(Table S5)**.

Since a poor R squared was obtained from the linear regression analysis, GLM analysis was subsequently conducted. Results from the GLM analysis reveal that after adjusting for all covariates, there is no significant correlation between PDQ-20 and TMT-A scores (β = -0.001, *p* = 0.751); however, age (β = -0.009, *p* < 0.001) and sex (β = -0.006, *p* < 0.013) are statistically significantly correlated with TMT-A performance **(Table 4)**. In contrast, after adjusting for covariates, there is a significant positive correlation between PDQ-20 and TMT-B scores (β = 0.003, *p* = 0.008) **(Table 5)**. Results also indicate that age (β = -0.009, *p* < 0.001), sex (β = -0.054, *p* = 0.010) and education (β = 0.052, p < 0.001) are statistically significantly correlated with TMT-B performance **(Table 5)**.

**Table 4.**
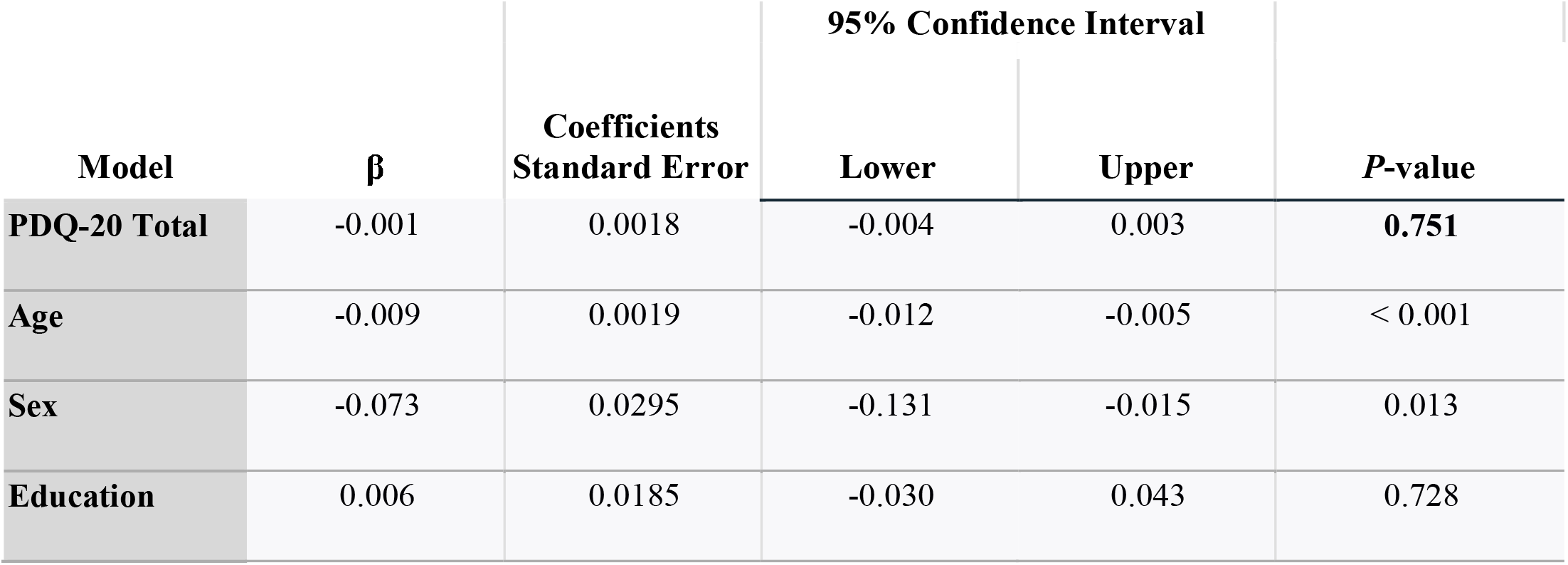
Generalized linear model for the relationship between PDQ-20 and TMT-A scores, after adjusting for age, sex, and education.

**Table 5.**
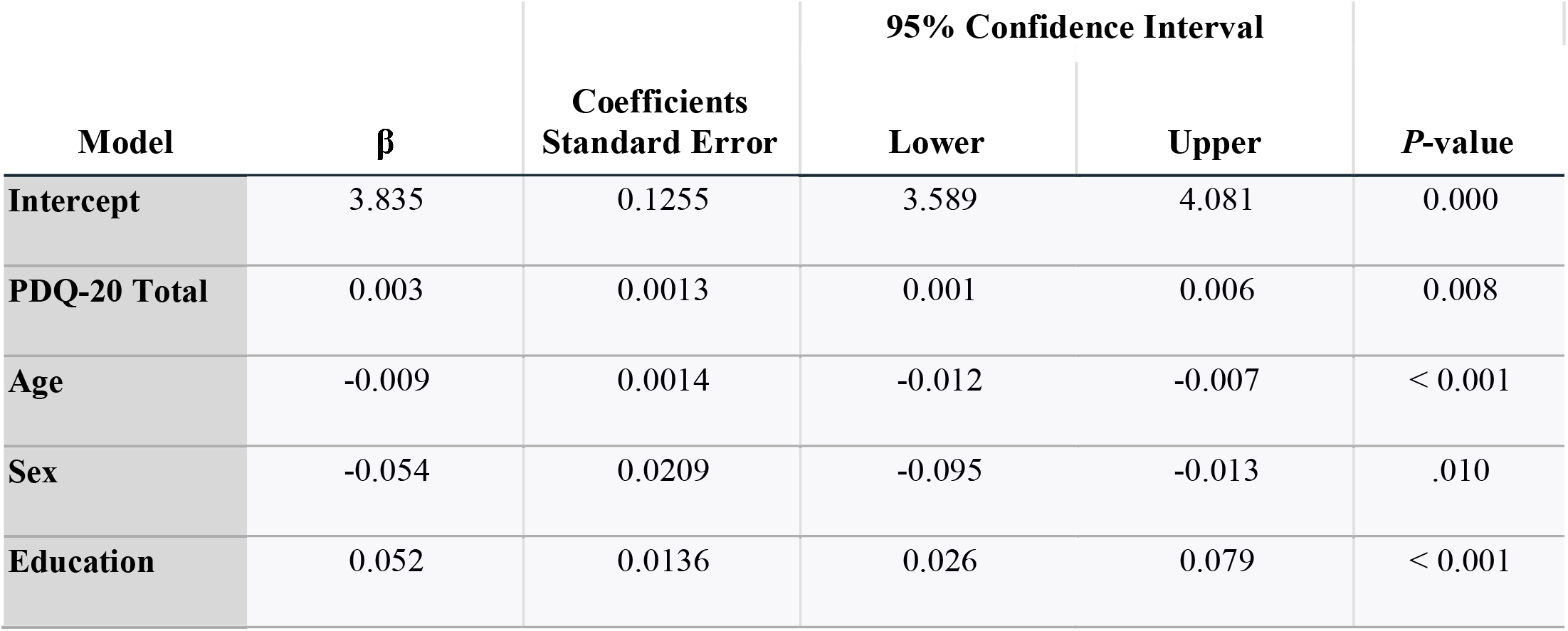
Generalized linear model for the relationship between PDQ-20 and TMT-B scores, after adjusting for age, sex, and education.

| Model | $\beta$ | Coefficients<br>Standard Error | 95% Confidence Interval | | *P*-value |
| --- | --- | --- | --- | --- | --- |
|  |  |  | Lower | Upper |  |
| **Intercept** | 3.835 | 0.1255 | 3.589 | 4.081 | 0.000 |
| **PDQ-20 Total** | 0.003 | 0.0013 | 0.001 | 0.006 | 0.008 |
| **Age** | -0.009 | 0.0014 | -0.012 | -0.007 | < 0.001 |
| **Sex** | -0.054 | 0.0209 | -0.095 | -0.013 | .010 |
| **Education** | 0.052 | 0.0136 | 0.026 | 0.079 | < 0.001 |

## DISCUSSION

Herein, we report an association between self-reported cognitive functions (i.e., attention/concentration, retrospective memory, prospective memory, and planning/organization) as measured by PDQ-20 and objective cognitive functions including processing speed, as measured by the DSST, and sets shifting, as measured by the TMT-B, in persons with PCC.^25^ In contrast, there was no significant association between self-reported cognitive functions and processing speed when measured with the TMT-A. Moreover, the correlation between subjective and objective cognitive functions persisted after controlling for the following covariates: age, sex, and education. Our results are in accordance with some but not all extant studies evaluating subjective and objective cognitive functions in persons with PCC.^1,26,27^ It is amply documented that across many medical disorders, a correlation between subjective and objective cognitive functions does not exist.^16,19^ Hitherto, however, there has not been sufficient understanding of whether significant correlations exist between subjective and objective cognitive functions in PCC.

Our results support the assumption that a significant percentage of persons with subjective cognitive complaints as part of PCC may have objectively verifiable objective cognitive problems. It cannot, however, be assumed that all individuals with subjective cognitive complaints have objective cognitive deficits and/or that the presence of subjective cognitive deficits in all cases is clinically relevant. Clinicians should evaluate affected persons for subjective cognitive complaints as part of their assessment of persons living with PCC. If present, clinicians should assist patients in modifying aspects that may improve overall cognitive abilities including, but not limited to, exercise, diet, discontinuation of alcohol and illicit substances, as well as pharmacologic agents with known anti-cognitive properties (e.g., benzodiazepines).^28–30^ Our results also inform future research endeavours that specifically aim to improve objective cognitive functions in persons living with PCC.

Several methodological limitations affect inferences and interpretations of our data. For example, this was a post-hoc analysis of data obtained as part of a primary study, and it was not pre-specified in the protocol that we would be exploring possible associations between objective and subjective cognitive functions. Although we ruled out other medical disorders as a primary reason for presentation, it is possible that other medical conditions patients had experienced in the past that could affect cognitive function were not mentioned. Furthermore, our self-reported measure of cognitive function in PCC was the PDQ-20 and objective cognitive measure was the DSST; it is possible that findings would be different if other cognitive measures or DSST/functional imaging analysis were included.^31^ In addition, our sample was heterogeneous with respect to acute COVID-19 severity, duration of PCC, number of prior COVID-19 infections, and number of prior vaccinations.

## CONCLUSION

Taken together, we observed a significant correlation between subjective and objective cognitive functions in persons with PCC. For individuals with PCC experience reporting subjective cognitive complaints, attention to interventions to reduce cognitive interference and potentially improve cognitive performance are encouraged. Future research studies should attempt to carefully ascertain which specific cognitive domains are most correlated with self-reported cognitive impairment.

## Data Availability

The data and research materials that support the findings of this study are available from the corresponding author, R.S.M, upon reasonable request and will be anonymized.

