## Supplemental Table 1 for "Subjective and Objective Measures of Cognitive Function are Correlated in Persons with Post-COVID-19 Condition: A Secondary Analysis of a Randomized Controlled Trial"

**SUPPLEMENTARY**

**Table S1.** Exclusion criteria

- Current symptoms were better explained by symptoms of major depressive disorder or bipolar disorder;
- Symptoms were fully explained by pre-existing conditions that may cause cognitive impairment or symptoms similar to those seen in PCC [e.g., attention deficit/hyperactivity disorder (ADHD), major neurocognitive disorder, schizophrenia, chronic fatigue syndrome (CFS)/encephalitis meningitis (EM), as assessed by Mini International Neuropsychiatric Interview (M.I.N.I.) 7.0.2];
- Known intolerance to vortioxetine and/or prior trial of vortioxetine with demonstrated inefficacy;
- Current alcohol and/or substance use disorder, as confirmed by the M.I.N.I. 7.0.2;
- Presence of comorbid psychiatric disorder that is a primary focus of clinical concern, as confirmed by the M.I.N.I. 7.0.2;
- Previous history of mania/hypomania;
- Taking medications approved and/or employed off-label for cognitive dysfunction (e.g., psychostimulants);
- Any medication for a general medical disorder that may affect cognitive function (as per clinical judgment);
- Use of benzodiazepines within 12 hours of cognitive assessments;
- Consumption of alcohol within eight hours of cognitive assessments;
- Any physical, cognitive, or language impairments sufficient to adversely affect data derived from cognitive assessments;
- Diagnosed reading disability or dyslexia;
- Clinically significant learning disorder by history;
- Treatment with electroconvulsive therapy (ECT) in the last 6 months;
- History of moderate or severe head trauma (e.g., loss of consciousness for > 1 hour), other neurological disorders, or unstable systemic medical diseases that are likely to affect the central nervous system (as per clinical judgment);
- Pregnant and/or breastfeeding; received investigational agents as part of a separate study within 30 days of the screening visit;
- Actively suicidal/presence of suicidal ideation or evaluated as being at suicide risk (as per clinical judgment);
- Currently receiving treatment with monoamine oxidase inhibitor (MAOI) antidepressants, antibiotics such as linezolid or intravenous methylene blue;
- Previous hypersensitivity reaction to vortioxetine or any components of the formulation;
- Previously reported angioedema in persons treated with vortioxetine;
- Serotonin syndrome;
- Abnormal bleeding;
- Angle closure glaucoma;
- Hyponatremia;
- Moderate hepatic impairment;
- Active seizure disorder/epilepsy that is not controlled by medication (as per clinical judgment);
- Presence of any unstable medical conditions;
- Inability to follow study procedures;
- And inability to give informed consent.

**Table S2.** Linear regression of PDQ-20 and combined DSST z-scores, after adjusting for age, sex, and education.

| **Model** | **Unstandardized B** | **Coefficients Standard Error** | **Standardized Coefficients Beta** | **95% Wald Confidence Interval** | |  | |
| --- | --- | --- | --- | --- | --- | --- | --- |
|  |  |  |  | **Lower** | **Upper** | **t** | ***P*-value** |
| **Constant** | 1.434 | 0.548 | n/a | 0.350 | 2.517 | 2.615 | 0.010 |
| **PDQ-20 Total** | -0.011 | 0.005 | -0.180 | -0.021 | -0.001 | -2.217 | 0.028 |
| **Age** | -0.022 | 0.007 | -0.264 | -0.035 | -0.009 | -3.313 | 0.001 |
| **Sex** | -0.005 | 0.093 | -0.004 | -0.189 | 0.180 | -0.050 | 0.960 |
| **Education** | 0.012 | 0.064 | 0.016 | -0.115 | 0.149 | 0.194 | 0.847 |

**a. Dependent Variable:** zdsst

**Table S3.** Linear regression of PDQ-20 and Pen/Paper DSST scores, after adjusting for age, sex, and education.

| **Model** | **Unstandardized B** | **Coefficients Standard Error** | **Standardized Coefficients Beta** | **95% Wald Confidence Interval** | |  | |
| --- | --- | --- | --- | --- | --- | --- | --- |
|  |  |  |  | **Lower** | **Upper** | **t** | ***P*-value** |
| **Constant** | 86.995 | 12.158 | n/a | 62.721 | 111.269 | 7.155 | < 0.001 |
| **PDQ-20 Total** | -0.190 | 112 | -0.216 | -0.413 | 0.033 | -1.697 | 0.094 |
| **Age** | -0.220 | 0.132 | -0.197 | -0.483 | 0.043 | -1.671 | 0.099 |
| **Sex** | -0.950 | 1.981 | -0.059 | -4.906 | 3.006 | -0.480 | -0.633 |
| **Education** | 0.639 | 1.466 | 0.053 | -2.288 | 3.565 | 0.436 | 0.664 |

**Table S4.** Linear regression of PDQ-20 and TMT-A, after adjusting for age, sex, and education.

| **Model** | **Unstandardized B** | **Coefficients Standard Error** | **Standardized Coefficients Beta** | **95% Wald Confidence Interval** | |  | |
| --- | --- | --- | --- | --- | --- | --- | --- |
|  |  |  |  | **Lower** | **Upper** | **t** | ***P*-value** |
| **Constant** | 28.913 | 20.920 | n/a | -12.868 | 70.694 | 1.382 | 0.172 |
| **PDQ-20 Total** | -0.005 | 0.192 | -0.004 | -0.388 | 0.378 | -0.026 | 0.979 |
| **Age** | 0.031 | 0.226 | 0.017 | -0.421 | 0.483 | 0.136 | 0.892 |
| **Sex** | -1.796 | 3.411 | -0.068 | -8.608 | 5.017 | -0.526 | 0.600 |
| **Education** | -0.176 | 2.534 | -0.009 | -5.237 | 4.884 | -0.070 | 0.945 |

**Table S5.** Linear regression of PDQ-20 and TMT-B, after adjusting for age, sex, and education.

| **Model** | **Unstandardized B** | **Coefficients Standard Error** | **Standardized Coefficients Beta** | **95% Wald Confidence Interval** | |  | |
| --- | --- | --- | --- | --- | --- | --- | --- |
|  |  |  |  | **Lower** | **Upper** | **t** | ***P*-value** |
| **Constant** | 48.226 | 39.559 | n/a | -30.778 | 127.231 | 1.219 | 0.227 |
| **PDQ-20 Total** | -0.037 | 0.363 | -0.014 | -0.761 | 0.688 | -0101 | 0.920 |
| **Age** | 0.031 | 0.428 | 0.009 | -0.824 | 0.886 | 0.072 | 0.943 |
| **Sex** | -0.0493 | 6.450 | -0.010 | -13.375 | 12.389 | -0.076 | 0.939 |
| **Education** | 1.781 | 4.791 | 0.048 | -7.788 | 11.350 | 0.372 | 0.711 |
